# A how-to-guide to building a robust SARS-CoV-2 testing program at a university-based health system

**DOI:** 10.1101/2020.06.03.20120832

**Authors:** Stephen D. Nimer, Jennifer Chapman, Lisa Reidy, Alvaro Alencar, Yanyun Wu, Sion Williams, Lazara Pagan, Lauren Gjolaj, Jessica MacIntyre, Melissa Triana, Barbara Vance, David Andrews, Yao-Shan Fan, Yi Zhou, Octavio Martinez, Monica Garcia-Buitrago, Carolyn Cray, Mustafa Tekin, Jacob McCauley, Philip Ruiz, Paola Pagan, Walter Lamar, Maritza Alencar, Daniel Bilbao, Silvia Prieto, Maritza Polania, Maritza Suarez, Melissa Lujardo, Gloria Campos, Michele Morris, Bhavarth Shukla, Alberto Caban-Martinez, Erin Kobetz, Dipen Parekh, Merce Jorda

## Abstract

When South Florida became a hotspot for COVID-19 disease in March 2020, we faced an urgent need to develop test capability to detect SARS-CoV-2 infection. We assembled a transdisciplinary team of knowledgeable and dedicated physicians, scientists, technologists and administrators, who rapidly built a multi-platform, PCR- and serology-based detection program, established drive-thru facilities and drafted and implemented guidelines that enabled efficient testing of our patients and employees. This process was extremely complex, due to the limited availability of needed reagents, but outreach to our research scientists and to multiple diagnostic laboratory companies and government officials enabled us to implement both FDA authorized and laboratory developed testing (LDT)-based testing protocols. We analyzed our workforce needs and created teams of appropriately skilled and certified workers, to safely process patient samples and conduct SARS-CoV-2 testing and contact tracing. We initiated smart test ordering, interfaced all testing platforms with our electronic medical record, and went from zero testing capacity, to testing hundreds of healthcare workers and patients daily, within three weeks. We believe our experience can inform the efforts of others, when faced with a crisis situation.

## Introduction

The optimal response of an academic health system to a global health crisis requires sustained coordination between academic faculty across many departments, and hospital and health system leadership, to develop the policies and procedures, communication strategies and teamwork needed for timely and effective decision-making. Novel crises place previously unidentifiable stressors across all aspects of an organization, and all weaknesses in the initial phases of a response must be rapidly identified and mitigated, through clear policies and procedures that are easy to disseminate to all involved.

The first cases of COVID-19 disease occurred in Wuhan, China, and were reported globally by Chinese health authorities on December 31, 2019. While the first confirmed COVID-19 case in the US was reported on January 21, 2020, the first case of COVID-19 disease in Florida was reported on March 1. At the time of this writing, Florida had 46,117 cases of COVID-19 disease, with 2096 deaths (as of May 20^th^, 2020)^1^.

The University of Miami Health system (UHealth), serves a four County catchment region with over 6 million inhabitants. We are the only University-based health system in the region, with three in-patient facilities and seven satellite locations. The University of Miami’s Miller School of Medicine (MSOM) employs 1100 physicians, with a departmental structure as well as institutes and centers, including the Hussman Institute for Human Genomics and Sylvester Comprehensive Cancer Center.

Our in-house COVID-19 testing efforts began March 30, 2020, when the Chief Clinical Officer and Interim Chief Operating Officer formed four leadership committees to address workforce management, clinical therapeutics, equipment and resources, and the *Testing Committee*, which was tasked with developing and implementing in house testing and algorithms for testing patients and employees.

Although our healthcare system has a longstanding and resilient emergency response infrastructure in place, based on extensive, annual preparations for the six month long hurricane season, preparation for the COVID-19 pandemic required the involvement of many additional personnel, with different skill sets, to address limited critical COVID-19 resources, develop and evolve testing capabilities and algorithms, while maintaining our standard of care for non-COVID-19 patients. Like other health systems, the widespread nature of the pandemic and the lethality of COVID-19 disease, led us to postpone or cancel non-essential patient visits and surgeries, and expand telemedicine capabilities. Social distancing, self-quarantine and other preventative measures were instituted, which helped “flatten the curve”.

Public health surveillance and testing strategies are the cornerstone for effective COVID-19 evaluation and management, so we assembled a team of pathologists, infectious disease experts, scientists, hospital administrators, laboratory technicians, public health practitioners, and others, who worked together to build a robust, multi-platform, PCR- and serology-based SARS-CoV-2 virus or antibody detection system. This system is being utilized to evaluate and monitor symptomatic patients, identify patients at risk, screen front line employees, and assess those with a history of close contact with infected individuals. This system is enabling us to identify contagious individuals and those likely to be immune. We have established contact tracing policies and procedures, implemented a COVID-19 convalescent plasma (CCP) program, and conducted IRB-approved clinical and translational research, including a program of community testing to define SARS-CoV-2 seroprevalence in our region.

## Developing in-house testing capability

Our COVID-19 testing was initially outsourced to commercial reference laboratories, but to meet our needs, we needed to rapidly develop in-house COVID-19 testing. We created three working groups that focused on implementing manual RT-PCR testing, automated/commercial PCR testing and serologic testing. Each group was charged with developing diversified testing platforms, in order to achieve the flexibility needed to deal with supply chain issues, including shortages or delays in the availability of reagents or instrumentation, and the changing regulatory environment, based on FDA or other agency regulations and state policies ^2^. The group was able to build a new molecular laboratory designed for manual RT-PCR-based testing within two weeks, getting reagents and instruments from the basic scientists on the medical and marine school campuses. We then utilized four separate CLIA certified clinical laboratories for insourcing SARS-CoV-2 testing, as shown in Figure 1. These test procedures were chosen based on the in vitro diagnostic Emergency Use Authorizations (EUAs) published on the FDA website ^3^ as well as available reagents and instrumentation. The testing platforms provide rapid turn-around time (TAT) and varying but extensive test capacity (Figure 1), which has been crucial to optimally manage specific groups of patients and increase capacity over time (Figure 2).

**Figure 1:**
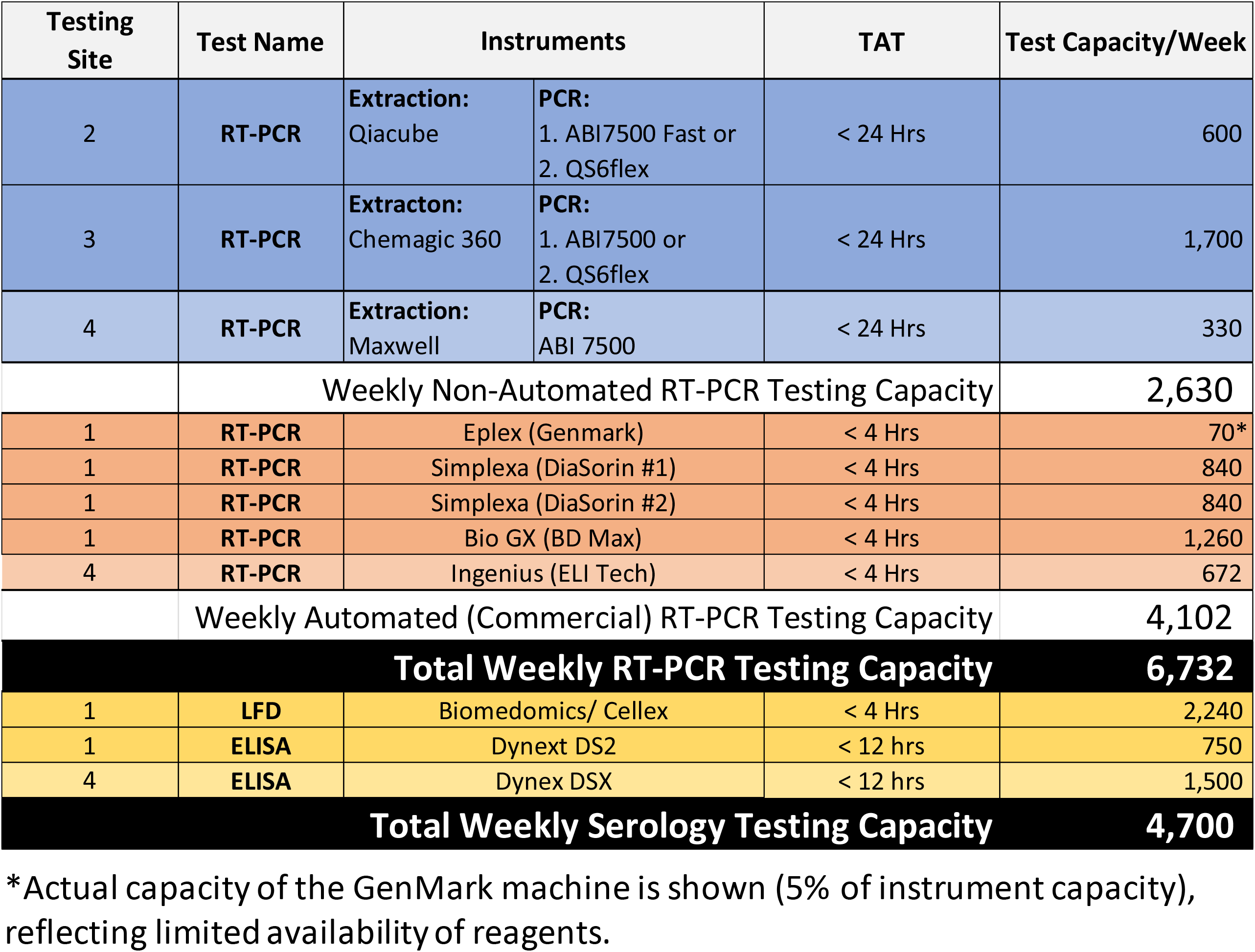
Testing platforms, turn-around times, and weekly test capacity. PCR and serologic testing platforms put in place, listing the instruments and workflow needed to perform any given test. Turnaround times (TAT) and test capacity per five-day workweek, based on reagents, machine capacity and personnel in place, are shown.

**FIGURE 2:**
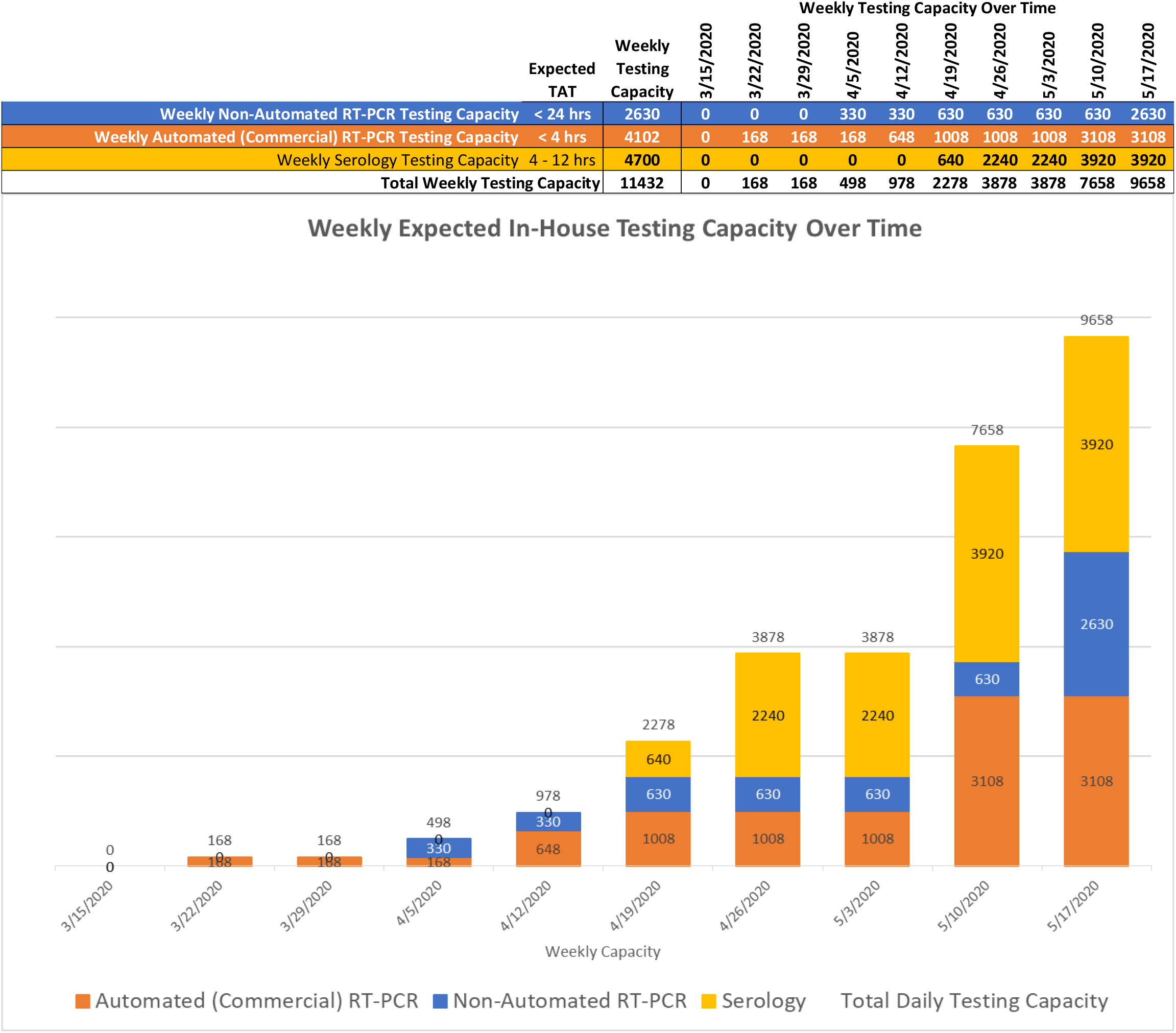
Weekly In-House Testing Capacity Over Time. Ramp up for weekly in-house COVID-19 testing shown, based on ordering additional reagents, hiring additional personnel, and the increased testing expected for patients and employees. Algorithm for determining how we can remap up PCR testing

Despite its limitations, serologic testing serves as an important adjunct to PCR testing, given the possible false negative rate of PCR-based testing for SARS-CoV-2 RNA^4^ and the need to detect evidence of prior infection, and possible viral immunity. Several types of immunoassays were evaluated, including lateral flow devices (LFD), enzyme-linked immunosorbent assays (ELISA), and chemiluminescence immunoassays (CLIA). Different tests detect different antibody isotypes (IgM, IgG, IgA, or total antibody levels), directed against different targets (e.g. the nucleocapsid, receptor binding domain and spike proteins) and with differing sensitivity^5-9^. Current analyses provide only a qualitative, positive or negative, result.

We validated the COVID-19 IgM/IgG Rapid Test kit, an LFD test from BioMedomics (Beckman-Dickinson) and the Epitope ELISA-based COVID-19 assay for IgM and IgG at our CLIA-certified high complexity laboratory. Using a venipuncture, serum or whole blood from in-patients was evaluated using the BioMedomics LFD test, which demonstrated 98%-100% specificity, and 71-73% sensitivity, respectively (Figure 3). This data is superior to the previous study from a larger validation cohort, which reported 89% sensitivity and 91% specificity. The ELISA Epitope validation demonstrated specificity of 95% for IgM and 90% for IgG, with an 85% specificity from the same patient samples. The BioMedomics test was initially evaluated as a EUA assay, pending FDA approval, so we implemented it as a high complexity Laboratory Developed Test (LDT). This required qualified laboratory personnel and a disclaimer in the result report stating that the test is not FDA approved. On May 7^th^, 2020, the FDA removed this test from the EUA pending list and hence we immediately ceased using it for clinical testing. Subsequently, we acquired the FDA EUA-approved SARS-COV-2 IgG/IgM Rapid Test by Cellex; we are in the process of validating this device in conjunction with the COVID-19 IgM/IgG test by Auto-bio Diagnostics (Hardy Diagnostics) to ensure that we have sufficient testing platforms and reagents on hand, for the ramp-up phase. As of May 20^th^, 2020, the Epitope IgG/IgM kit is still pending FDA-EUA approval.

**FIGURE 3:**
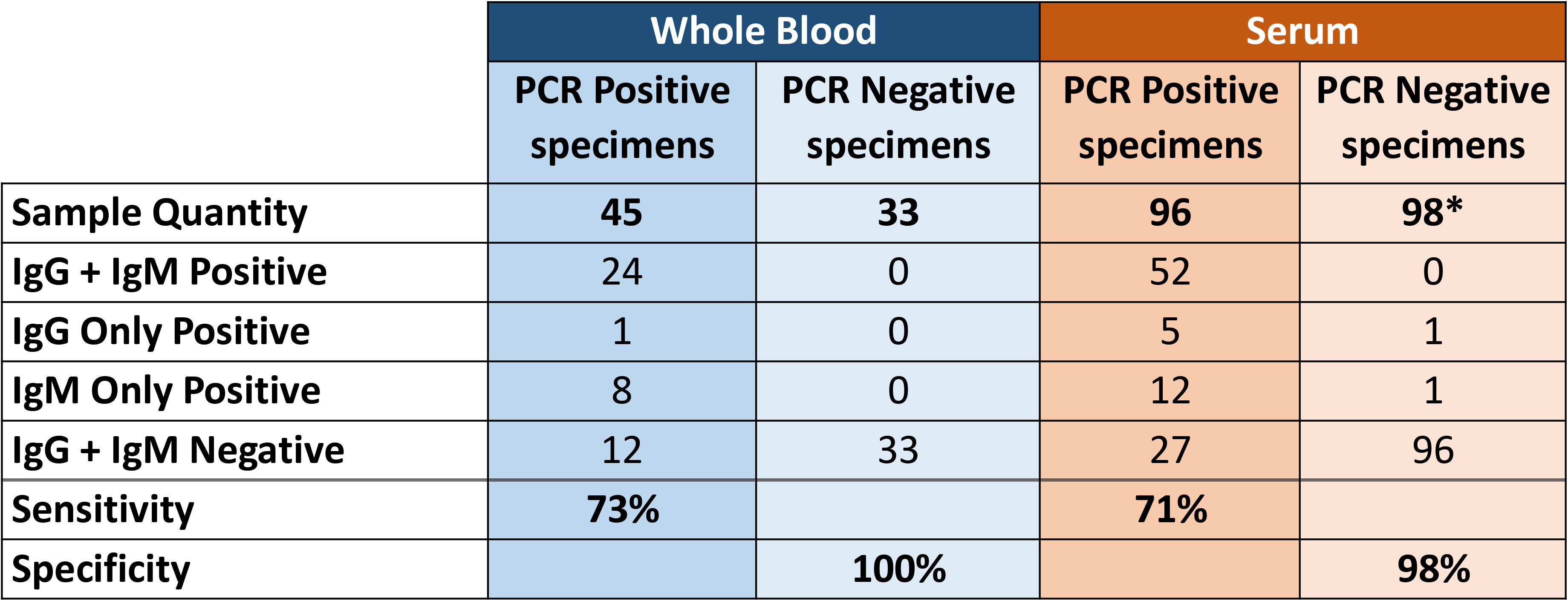
Validation of the Biomedomics LFD Serologic Test. Validation results of the BioMedomics lateral flow device (LFD) serologic test. Testing of whole blood and serum was conducted and compared to PCR results. Overall 272 samples were tested; the sensitivity was 72-73% while the specificity was 97-100%. The validation cohort included specimens collected at a variable time from the onset of symptoms or the PCR test was positive.

We continue to encounter problems with reagent availability, especially those that have received FDA EUA approval, despite having assembled a team of project managers, industrial engineers, administrators, physicians and scientists, who along with the institution’s supply chain leaders and COVID-19 command center institutional team assure all requests for instruments and supplies are coordinated with industry and government agencies. We have also created interactive Gantt Charts that allow us to track availability of supplies on a daily basis and assess staffing needs.

We created a data mining and reporting team, to ensure that institutional, state and federal reporting data requirements are met. Key information reported included the number of tests performed each day, the percentage of positive results, up to date turn-around-times for all laboratories, volume of orders by site and department, number of pending results, and number of rejected, not performed or canceled tests, among others. We leveraged the electronic medical record and laboratory information system to capture information on the performing laboratory, the methodology and instrumentation used, the reference range, CPT codes for billing, results and interpretation, and the required disclaimers stating the FDA status of EUAs or LDTs and the limitations of the tests^10-12^.

## Community-based surveillance and testing initiatives

During the initial days of the pandemic, we worked with State, County and City officials and agencies to support the establishment of community testing initiatives, helping establish best practices for the first drive thru testing site. Committee members toured the sites under development and provided nursing and other personnel to support the appointment scheduling lines. Working with Miami-Dade County (MDC) officials, we also established a surveillance program (see below) built upon random testing of cross sections of the county’s 2.8 million population, using Biomedomics anti-SARS-CoV-2 antibody detection kits. We also provided surveillance and testing support to our first responders.

## Employee Testing-establishing a robust program

### Call center

A multi-disciplinary group involving employee health, infection control, public health experts and human resources devised a comprehensive algorithm for assessing returning travelers, as well as potential exposures within the community or the health care system. Our standard employee health processes and infrastructure were insufficient to manage the myriad of issues that the COVID-19 pandemic raised, requiring the development of a robust Employee Health call center capable of rapidly triaging the hundreds of calls received daily. We created a dedicated, centralized phone line, that handled calls from employees to report positive test results, exposure to or potential symptoms of COVID-19, and was staffed by APRNs, RNs and call center personnel. We established training procedures and algorithms that addressed employee testing, quarantine and return to work policies to maintain a safe, essential healthcare workforce. Then, we began to order, schedule and coordinate appointments for testing, providing testing instructions, communicate test results and provide directions on employee return to work dispositions, including the need for quarantine. An Epidemiology team was formed to conduct contact tracing on individuals with positive results and those who were presumptively positive but did not meet testing criteria (e.g. employees who were able to complete work from home). These practices were approved by our HIPPA, Privacy and Risk Management offices to ensure that employee data was appropriately managed. Later, we used the expertise of this staff to support other rapidly evolving needs, such as the testing and processing of potential donors for our COVID-19 convalescent plasma infusion research program. As of May 15, 2020, the call center had received 4925 calls, and ordered 881 tests.

### Drive Thru Testing Initiatives

To support the critical need for testing employees for COVID 19, we established a University-operated drive through test site within 72 operational hours, capable of performing 84 nasopharyngeal swab tests per day, that uses a paperless check-in process that includes a 4-6 question questionnaire. The site was created to comply with all Authority Having Jurisdiction (AHJ) standards and use best practices to maintain staff and patient safety.

### Employee contact tracing

We also established a formal contact tracing program called UM Tracking, Resources, and Assessment of COVID-19 Epidemiology Program (U-TRACE), which engaged faculty with expertise in epidemiology, occupational health and safety, nursing, informatics, environmental exposure assessment, and public health education (Figure 4). To implement U-TRACE, we conducted a needs assessment of key stakeholder groups, designed the contact tracing workflow and data collection instruments, implemented a relational database to monitor and track COVID-19 cases, and adapted the program to federal, state and local University/UHealth policy. This supported the coordination, tracking and education of COVID-19 contact, presumptive and confirmed cases among faculty, staff and trainees. We used REDCap, a secure, HIPAA compliant web-based application, to establish a relational database for our contact tracing, that allowed us to enter, review or track COVID-19 persons under investigation (PUIs), as well as COVID-19 cases among university and hospital employees. We established an employee hotline (XXX-XXX-TEST) to assist in contact tracing, which provided an initial clinical assessment of symptoms and triaged employees based on a risk assessment and separate return-to-work flow diagram. Employees flagged for contact tracing were referred to the U-TRACE team by a telephone encounter note available in the Electronic Health Record, by telephone or email. For each co-worker identified by the employee (“patient zero”), the U-TRACE made three attempts to contact the co-worker for referral and screening to the main employee hotline. To date, the U-TRACE program has completed 1,681 employee encounters of which 357 resulted in contact tracing due to COVID-19 presumed or confirmed positivity, or positive COVID-19 symptoms. Approximately 81% of the employees that required contact tracing provided permission to let their co-workers know they were being evaluated as being potentially SARS-CoV-2 infected.

**FIGURE 4:**
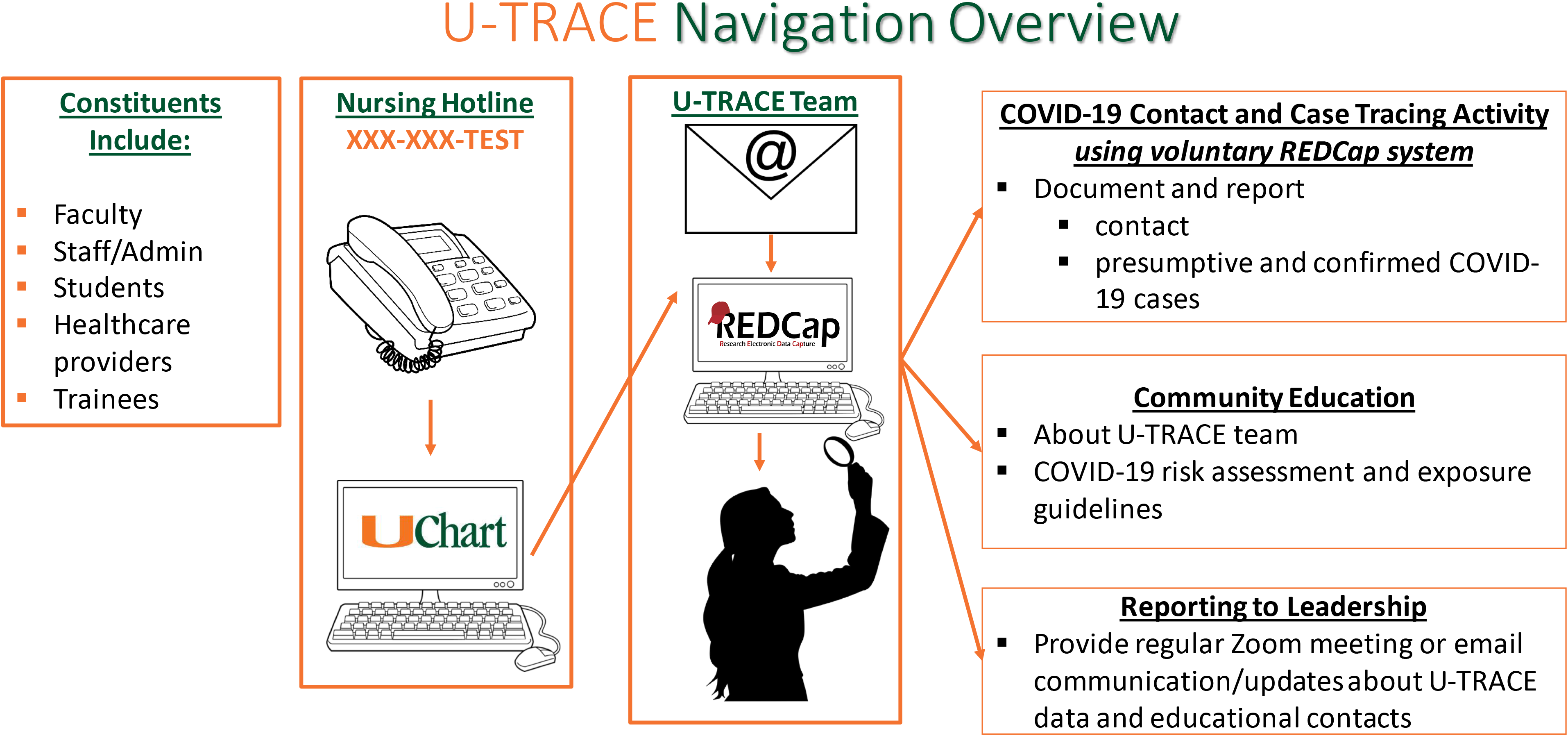
Contact Tracing Program. Contact tracing navigation is shown for faculty, staff, students, health care workers and trainees.

### Employee serologic testing initiative

Healthcare workers (HCWs) are at high risk for SARS-CoV-2 infection and, especially if asymptomatic, could unknowingly transmit the virus to colleagues and/or patients. As part of our back-to-work strategic planning, to identify asymptomatic but possibly infected HCWs, we examined the utility of serologic testing, to be reflexed to PCR testing where appropriate (Figure 5, right panel), using an IRB-approved research protocol and the BioMedomics Rapid IgM/IgG test that we internally validated. We enrolled 500 asymptomatic HCWs (out of over 1600 HCW volunteers) who were either working in clinic areas or in the hospital, and thus had potential exposure to COVID patient/samples, over a two-three week period. Results of this study are currently being analyzed.

**FIGURE 5:**
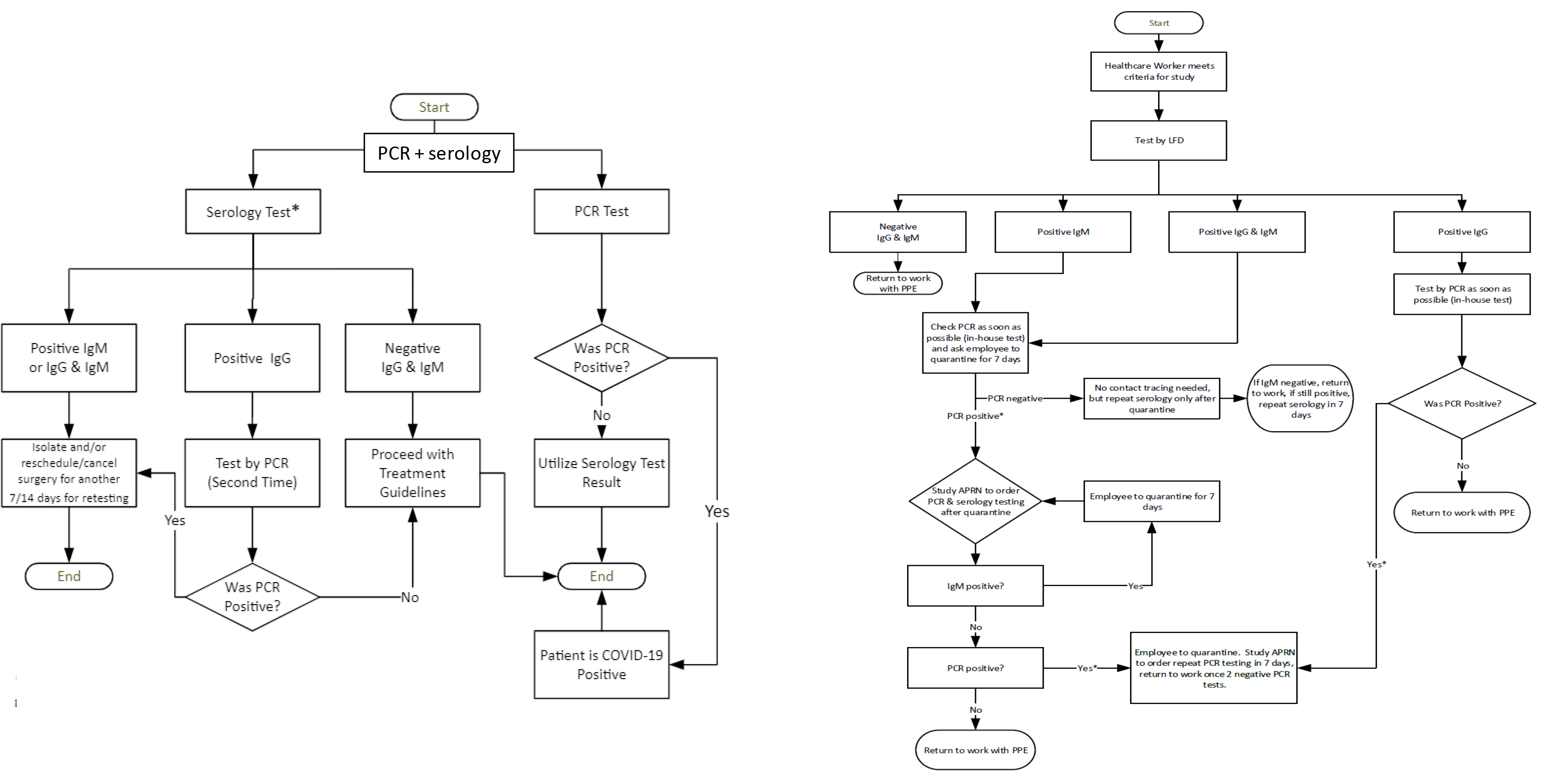
Algorithms for Asymptomatic Patients Undergoing High Risk Procedures and Asymptomatic Healthcare Workers. Testing algorithms for (left) asymptomatic patients undergoing high-risk procedures, involving a combination of PCR and serologic testing, and (right) asymptomatic health care workers, based on an IRB approved study of 500 employees, using serologic testing as the primary testing strategy, with reflex RT-PCR testing to determine whether employees with a positive serologic test have detectable SARS-CoV-2 virus.

## Patient-testing algorithms

Our initial patient testing algorithms were based on the three-tiered CDC priority system for testing patients with suspected SARS-COV-2 infection^13^. Limited access to required personal protective equipment (PPE) ^14^, nasopharyngeal (NP) swabs and varying turnaround times were the greatest constraints, thus, besides patient acuity, we incorporated the impact of utilization of scarce resources into our algorithm. Because our testing capabilities were rapidly expanding, our triage system needed to be flexible, so we also defined three phases of our SARS-CoV-2 testing program, with the first phase being the roll out phase. We did not define the second and third phases immediately, nor provide a date or criteria that we would use to announce movement from one phase to another. The rollout phase focused on top priority cases including those being admitted to the hospital, and those patients who required emergency interventions or a high-risk procedure (Figure 5, left panel). Our intent was to later include high-risk populations in the testing program, such as long-term facility residents and symptomatic patients older than 65 or with underlying health conditions. A team of clinicians and epidemiologists were involved in drafting these guidelines and in reviewing individual cases to approve prompt testing when a high likelihood of infection was suspected. Soon after we rolled out our patient testing algorithms, we began testing all admissions through the emergency department, and all elective admissions for cancer care 48-72 hours prior to admission.

## Established and Evolving COVID-19 Research efforts

### Community-based testing program

Working with key administrators from the MDC Mayor’s Office, and call center leadership from Florida Power and Light (FPL), we developed a community testing program called The Surveillance Project Assessing Risk and Knowledge of Coronavirus (SPARK-C), to approximate the seroprevalence of SARS-CoV-2 antibodies in MDC residents, determining the proportion of individuals infected, as well as their age, gender, and racial/ethnic distribution, their most common symptoms and the fraction of asymptomatic infections. The FPL team randomly selected phone numbers to cover the geographic breadth of MDC, played an automated/commercial voice recording of the County Mayor, encouraging people answering the phone to participate, and then antibody screening was conducted at a testing site nearest their home, typically a local park or library, after they signed an informed consent form. Serologic testing was conducted using the Biomedomics serologic test. However, once the FDA removed this test from the EUA list, the SPARK-C program was temporarily paused to reconsider the best strategy for implementation.

### Support for a convalescent plasma infusion program

We have established a COVID-19 convalescent plasma (CCP) infusion program, as case series from China have shown promising results using convalescent plasma to treat patients with severe COVID-19 ^15-17^. With our efforts in ramping up COVID-19 testing capability both for molecular testing and for serological testing, we established the following process to support CCP collection: 1. Created pathway to perform COVID-19 testing for potential CCP donors; 2. Make COVID-19 testing results available for potential CCP donors; 3. Consent patients who require COVID-19 testing for future contact for potential CCP donation. To date, we have collected convalescent plasma, with assistance from OneBlood, from dozens of individuals.

### Biospecimen acquisition and analyses

To promote multidisciplinary COVID-19 research, we have developed a COVID-19 clinically annotated biobank which contains consented and de-identified samples, including blood (serum, plasma, buffy coat samples), and cytology fluids, saliva, urine, fine needle aspirates, and surgical or autopsy tissue samples. Access to these materials will foster laboratory research, support novel clinical trials and ideally generate data for NIH and other grant proposals.

## Conclusions

We activated a health system crisis response team in early March to tackle the unique challenges associated with the COVID-19 pandemic, including supply limitations (e.g. nasopharyngeal swabs, collection kits, transport media and regent kits), that were severely limiting our response, including our ability to perform PCR testing to detect SARS-CoV-2 virus. Necessary, but otherwise non-descript, products such as nasopharyngeal swabs, collection kits, transport media and reagent kits were in acute shortage. Commercial laboratories and state health departments faced similar shortages and challenges and soon testing turnaround times ballooned to more than 5-7 days. We quickly realized the pitfalls of relying heavily on any single testing platform or vendor and created a testing committee charged with overseeing every aspect of testing. We pulled together resources, processes, expertise, reagents, machines, decision-making tools from across the entire campus, including the academic leaders, the laboratory researchers, the operational leaders, and the procurement teams. Together, we assembled numerous testing platforms, managed reagent inventory, communicated with external vendors and developed testing algorithms for our patients and our employees. The strength of the university’s research enterprise enabled us to rapidly build up in-house testing capabilities, including serologic testing, which we ordered in bulk during the early phase of our testing efforts, with goal of subsequently validating the assays in house.

What is most evident from the current COVID-19 environment is the importance of planning, effective resourcing, nimble regulatory bodies and clinical scientific rigor. To achieve these goals, we developed a leadership structure, assigned tasks to key individuals, evolved effective twice-daily communications via teleconferencing and aligned accountability with responsibility. This was done in a rapidly changing environment, with different authorizations and approvals (mostly EUAs) coming from the FDA several times each week. Given the lead-time to develop new testing platforms, we had to commit to certain workflows, even though improvements came along that necessitated changes in the reagents used, the instruments utilized, or the body fluids subjected to analysis.

The role of accessible diagnostics is ‘front and center’ in this crisis – not only for knowing whether someone has the virus but also whether they may be immune to subsequent infection but also be a potential donor for our CCP donation program.

We hope our experience in bringing together health care personnel from the health system, the university’s research community, and its administrative leadership, can instruct other health care systems rapidly ramp up their ability to test for pathogens and implement health care policies that enable them to effectively respond to a health crisis.

## Data Availability

The data in this manuscript has not been published elsewhere. Primary data will be made available to anyone who wishes to review it.

## Acknowledgements

We thank the many dedicated healthcare personnel who have worked under difficult conditions to keep our patients and employees safe. We also thank many members of UM, UHealth and our community for assisting in establishing and maintaining our COVID-19 testing and surveillance program.

## Authors’ contributions

SDN, MJ and DP contributed to the conceptualization of the manuscript, SDN and MJ to the organization, editing and final drafting of the document. JC, LR, AA, YW, SW, LP, LG, MGB, CC, JM, MS, GC, MM, BH, ACM, EK, contributed to operational and technical aspects of the work described in this report, to drafting the document, and final approval. DA, YSF, YZ, OM, MT, BV, MT, JM, PR, PP, SP, MP, ML, contributed to operational and technical aspects of the work described in this report and to final approval. All authors agree to be held accountable for all aspects of the work related to its accuracy and integrity.

## Institution and Ethics approval and informed consent

The University of Miami’s Institutional Review Board gave approval for the seroprevalence study in asymptomatic healthcare workers.

## Disclaimers

None

